# Towards Metacognitive Clinical Reasoning: Benchmarking MD-PIE Against State-of-the-Art LLMs in Medical Decision-Making

**DOI:** 10.1101/2025.01.28.25321282

**Authors:** Yasma Esteitieh, Shaurjya Mandal, George Laliotis

## Abstract

The ability of large language models (LLMs) to perform clinical reasoning and cognitive tasks within medicine remains a critical measure of their overall capabilities in decision-making, with significant implications for patient outcomes and healthcare efficiency. Current AI models often face limitations in real-world clinical environments, including variability in performance, a lack of domain-specific knowledge, and black-box reasoning processes. In this study, we introduce a novel PIE framework, named MD-PIE, which emulates cognitive and reasoning abilities in medical reasoning and decision-making. We benchmark our framework and baseline methods both quantitatively and qualitatively using state-of-the-art LLMs, comparing them against OpenAI’s o1, Gemini 2.0 Flash Thinking, and DeepSeek V3 across diverse benchmarks. Our results demonstrate that MD-PIE surpasses existing models in differential diagnosis and reasoning accuracy across diverse medical benchmarks. This study underscores its potential to improve clinical decision-making through adaptive and collaborative design. Future research should focus on larger benchmarks and real-world validation to confirm its reliability and effectiveness in varied clinical scenarios.

## Introduction

The integration of artificial intelligence (AI) in healthcare has significantly advanced over recent decades, with Large Language Models (LLMs) emerging as a cornerstone in enhancing medical decision-making processes (1–4). Clinical reasoning is a multifaceted and iterative process that encompasses the continuous interpretation and synthesis of patient data, the assimilation of new information, the generation of differential diagnoses, and the formulation of effective management plans (5). Traditionally, these tasks have relied on the cognitive expertise and collaborative efforts of multidisciplinary teams.

Recently, LLM-based clinical decision support systems (CDSS) have shown great promise in aiding diagnostic and treatment processes by providing impactful recommendations (6–14). These models excel at processing large volumes of biomedical data and have even surpassed human clinicians on various standardized benchmarks. However, most evaluations occur in static, task-specific setups, failing to address the demands of real-world clinical scenarios, which require dynamic, multi-step reasoning that is both explainable and iterative. Physicians must continually adjust decisions based on new and diverse data, refine differential diagnoses, and manage high-stakes treatment decisions under uncertainty (15). This gap in adaptability, higher-order reasoning, and domain-specific knowledge undermines physicians’ trust in these systems, limiting their practical utility (16). While recent advancements, such as OpenAI’s o1-preview, show promise in diagnostic reasoning and probabilistic assessments, these limitations hinder the full realization of LLMs’ potential to enhance clinical decision-making (17).

Our study addresses current limitations in medical decision-making and reasoning by benchmarking MD-PIE, a novel LLM framework, against state-of-the-art AI models (9). MD-PIE utilizes a dynamic, multi-agent collaboration methodology designed to handle realtime, complex, multi-step reasoning and adapt to the challenges of medical tasks (9–10). We evaluate its performance against leading LLMs, including o1, Gemini 2.0 Flash Thinking, and DeepSeek V3, across diverse clinical reasoning benchmarks. The results demonstrate that MD-PIE outperforms these models, offering a highly accurate, efficient, and user-friendly solution with significant potential to enhance clinical reasoning and reduce diagnostic errors, reinforcing the promise of AI in medical practice. Our findings highlight the importance of LLMs in improving diagnostic accuracy, efficiency, and the overall quality of healthcare services. This study contributes to advancing the understanding of how adaptive and collaborative AI systems can emulate and enhance clinical workflows, setting a new standard for AI-assisted medical decision-making.

## Methods

### Formulation of the Problem of Inclusion-Exclusion: MD-PIE

The diagnostic process can be conceptualized as a structured approach to systematically gather and interpret patient information, with the ultimate aim of identifying the most probable diagnosis. In this study, we propose modeling the diagnostic workflow as a Problem of Inclusion-Exclusion (PIE). Within this framework, a large language model (LLM) iteratively solicits symptom information, refining its diagnostic hypotheses by incorporating findings consistent with the patient’s presentation and excluding those that are contradictory or irrelevant. This approach is designed to achieve three primary objectives: (1) High Diagnostic Accuracy (*α*): Ensuring that the final diagnosis aligns with clinical evidence; (2) Low Error Rate (*ϵ*): Minimizing the likelihood of misdiagnoses or missed conditions; and (3) Reduced Human Intervention (*β*): Streamlining clinical workflows by automating routine elements of the diagnostic process.

### Bridging to Enhance Diagnostic Accuracy

Bridging refers to the process of augmenting a large language model (LLM) with additional, contextually relevant information obtained from external knowledge bases, domain experts, or specialized computational models. This augmentation is critical for guiding the diagnostic reasoning process toward well-supported and accurate conclusions, especially in complex or overlapping clinical scenarios where the integration of cross-domain data is essential. By compensating for the LLM’s inherent limitations in synthesizing multifaceted information, bridging helps prevent several key issues. These include diagnostic hallucinations, where the model generates non-existent conditions or symptoms; repetitive queries, which involve inefficient loops of redundant questioning that do not contribute to diagnostic clarity; and the omission of subtle clinical signals, where less obvious but critical findings are overlooked. By incorporating diverse and complementary sources of information, bridging strengthens the robustness and reliability of the diagnostic process, ensuring more comprehensive and clinically relevant outcomes.

### Symptom Set for Each Condition

Let *C* = *{c*_1_, *c*_2_, …, *c*_*m*_*}* be a set of possible conditions. Each condition *c*_*i*_ is associated with a set of symptoms *S*_*i*_ *⊆ U*, where

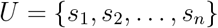

is the universal set of symptoms across all conditions. While many conditions in *C* may share overlapping symptoms, each one also has a unique combination of signs or clinical markers. The overarching objective is to identify the correct condition *c*^*\**^*∈ C* by asking the user (i.e., the patient) about the presence or absence of symptoms in *U*.

### Query Process

During a diagnostic conversation, let *Q⊆ U* denote the set of symptoms the LLM has already asked about, and let

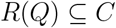

represent the set of possible conditions based on the user’s responses thus far. Initially, *R*(*Q*) = *C*. Each new symptom query aims to reduce the size of *R*(*Q*). Formally, when the LLM queries a new symptom *s*_*k*_, the system updates *R*(*Q*) as follows:

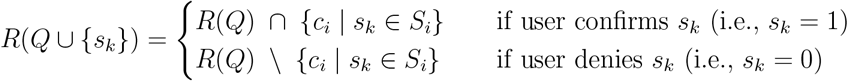

Thus, each user response either *includes* conditions in which the symptom is known to manifest or *excludes* those conditions if the user denies having that symptom.

### Query Selection

To minimize the number of questions while maintaining high accuracy, the LLM selects the *next best symptom s*_*k*_ *∈ U\ Q* by maximizing an *information gain* function combined with a *set-balance* measure. This approach ensures that queries both reduce uncertainty and partition the remaining condition set more evenly. Specifically, we define:

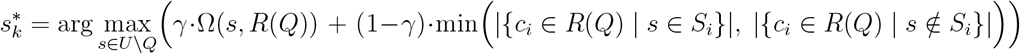

where

- *γ* is a weighting factor (0.5 by default) that trades off *information gain* vs. *set balancing*.
- Ω(*s, R*(*Q*)) is the *information gain* function, computed as:

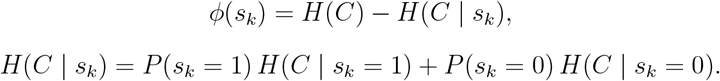 Here, *H*(*C*) is the entropy of the set of conditions, and *P* (*s*_*k*_ = *r*) are the probabilities for each potential user response *r ∈ {*0, 1*}*.
- min(|*{c*_*i*_ | *s ∈ S*_*i*_*}*|, |*{c*_*i*_ | *s ∉ S*_*i*_*}*|) aims to *balance* the partition of conditions, mitigating extreme splits where one response leads to a very small or large subset of possible diagnoses.

### Utility Function

To further incorporate practical objectives such as accuracy (*α*), error rate (*ϵ*), and reduced human intervention (*β*), we refine the query selection rule with a utility function:

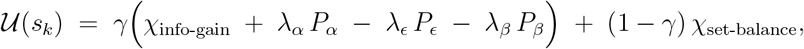

where:

- *P*_metric_ = *P* (metric | *s*_*k*_) are the probabilities that querying *s*_*k*_ improves accuracy, reduces errors, or diminishes the need for human guidance (e.g., bridging with a physician).
- *λ*_metric_ are importance weights for each of these metrics, typically set to 0.33 for equal consideration.

### Convergence Requirement

The iterative query process continues until only one possible condition remains, i.e., |*R*(*Q*) | = 1. At this point, the model has converged on the most likely condition *c*^*\**^. The overarching goal is to find the minimal set of queries *Q ⊆ U* that yields this convergence:

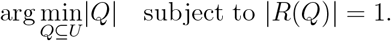

Although the above framework is readily generalizable to medical conditions of varying complexity, we note that practical implementations often balance thoroughness (e.g., asking many questions) against user fatigue.

### Constructing the Inverse Prompt

*Inverse prompting* refers to the construction of a *system prompt* that directly seeds the LLM with a set of relevant symptoms for the condition space under consideration. By doing so, we ensure that the LLM does not have to *discover* these symptoms from scratch—an action prone to hallucination or inaccurate assumptions. Instead, the LLM is primed to *actively ask* about these predefined symptoms in a systematic manner.

Formally, given conditions *C*_1_, *C*_2_, …, *C*_*N*_ each with unique symptom subsets, we can create a “composite” or synthetic set of symptoms that represents a diverse range of clinical presentations. These symptoms are used to *warm-start* the model, ensuring it queries logically and avoids generating extraneous or irrelevant symptoms. This prompt design is particularly beneficial in scenarios where an accurate, structured dialogue about known symptoms is paramount to effective diagnosis.

### Multiagent Collaboration for Specialist Input

In more complex clinical scenarios—such as when a patient’s symptoms overlap with multiple specialties (e.g., neurology, orthopedics, cardiology)—the PIE framework can be enhanced through a *multiagent* approach. Specifically: Primary Diagnostic Agent; the main LLM, which orchestrates the query process and performs the core inclusion-exclusion reasoning, Specialist Agents; one or more domain-specific agents (e.g., a neurology-focused agent, an endocrinology-focused agent) that the primary LLM can query when it identifies domain-specific complexities. This collaboration is initiated if the utility function *𝒰* (*s*_*k*_) indicates that bridging from an expert is likely to reduce error (*P*_*ϵ*_) or improve accuracy (*P*_*α*_).

At runtime, if the LLM encounters ambiguous or cross-domain symptoms, it dispatches a query to the relevant specialist agent. The response from the specialist agent is incorporated back into the bridging mechanism to guide subsequent questions and reduce diagnostic uncertainty. This multiagent system thus extends the PIE methodology by adding an extra layer of domain-expert verification, significantly augmenting the reliability and interpretability of the final diagnosis.

### User Interaction Logic

With the system primed via an inverse prompt and specialists on standby for bridging, the patient (user) initiates a conversation by describing one or more symptoms. Let *P* be the set of symptoms actually experienced and mentioned by the patient, with *P ⊆ U*. As the consultation progresses:

1. The model asks clarifying questions (drawn from *U*) to further narrow down the remaining set of conditions *R*(*Q*).
2. If encountering an intricate symptom, the model engages the appropriate specialist agent for confirmation or deeper insight.
3. The process repeats until |*R*(*Q*) | = 1 or until the probability distribution over *R*(*Q*) indicates a single highly probable condition.

This interactive process ensures that the patient’s clinical picture is accurately captured while reducing the risk of irrelevant or redundant inquiries. The resulting conversation— encompassing both the patient’s responses and the model’s rationale—can then be forwarded to a healthcare practitioner for validation and final decision-making.

We have presented a PIE-based methodology designed to enhance medical diagnosis with LLMs. We iteratively reduce the differential diagnosis space by framing symptom queries through an inclusion-exclusion lens. Integrating bridging and multiagent collaboration ensures that domain complexities and cross-specialty knowledge gaps are adequately addressed, thus minimizing errors and improving the clarity of diagnostic recommendations. Although this is an initial framework, we envision that future agent-based solvers—equipped with real-time entropy computations and refined utility functions—will further reduce query complexity, accelerate convergence, and improve patient satisfaction in clinical and telehealth environments.

## Quantitative Benchmarking Methodology

### MedQA Dataset Overview

For our experiments, we utilized the English questions from the MedQA dataset, a widely-used medical-question answering benchmark that contains clinically oriented queries. The dataset simulates a variety of scenarios similar to those found in medical board examinations, thereby providing a robust evaluation of a model’s diagnostic reasoning capabilities. In particular, the questions challenge the model to interpret clinical vignettes, laboratory results, and imaging findings in order to arrive at the most probable diagnosis or management plan.

### Experimental Setup

We conducted three separate experiments to evaluate how different reasoning strategies affect model performance on MedQA. In each experiment, we tested the following models: OpenAI o1: A proprietary model with a strong baseline in natural language comprehension; Gemini 2.0 Flash Thinking: A new generation LLM that emphasizes rapid inference for medical reasoning; DeepSeek V3: A specialized healthcare-focused model trained extensively on medical texts and case studies. The primary metric employed in all experiments was accuracy (i.e., the proportion of questions for which the model selected the correct answer).

### Experiment 1: Zero-Shot Reasoning

In the first experiment, all three models were presented with the raw MedQA questions without any additional instructions or context on problem-solving strategy. This zero-shot approach tests how well each model can reason directly from the clinical vignette or question prompt.

### Experiment 2: Instruction-Guided Reasoning

In the second experiment, we introduced a brief instruction prompt before each question, guiding the models to outline their thought processes and step-by-step reasoning. This additional contextualization aimed to improve model performance by clarifying the logical steps required for solving a clinical problem.

### Experiment 3: PIE-Integrated Reasoning

Finally, we integrated the Problem of Inclusion-Exclusion (PIE) formulation into each model. As described in the previous section, PIE systematically queries the user about relevant symptoms to include or exclude conditions. **OpenAI o1 + PIE**: Incorporates a structured, symptom-based interrogation loop. **Gemini 2.0 + PIE**: Augments its Flash Thinking method with iterative queries and updates. **DeepSeek v3 + PIE**: Combines its medical domain expertise with the PIE querying strategy.

## Qualitative Benchmarking Methodology

### Models

The following models were evaluated for qualitative performance: o1 Full Version (“o1-full version-2024-12-05”) accessed online; DeepSeek V3 (“DeepSeek-2024-12-26”) accessed online; Gemini 2.0 Flash Thinking (“Gemini 2.0 Flash Thinking-2024-12-19”) accessed online. MD-PIE was tested with GPT-4o as the base LLM.

### Experiment 1: NEJM CPC - Differential Diagnosis Quality

We gathered five diagnostic case challenges from March 2024 to November 2024, excluding the section labeled “Differential Diagnosis.” To predict differential diagnoses, we adapted a prompt initially used in a previous GPT-4 study (Supplement 1A, supplemental material is available upon request).

Our primary focus was on the quality of the model’s and the physician-solvers’ differential diagnoses. Each differential diagnosis output was evaluated independently by a single specialist physician (G.L.) using the previously developed Bond Score system (18). Under this scoring system, values range from 0 to 5, where a score of 5 indicates that the differential list contains the exact target diagnosis, whereas a score of 0 means that none of the listed diagnoses come close to the correct one (Supplement 1B).

### Statistical Analysis

The comparison of the performance on diagnosis quality of MD-PIE, o1, Gemini 2.0 Flash Thinking, DeepSeek V3, and Physicians on NEJM CPCs was evaluated using a McNemar’s test based on their ability to identify an exact or very close diagnosis (i.e., Bond score 4/5 or 5/5) versus not (i.e., Bond score 0/5, 2/5, or 3/5). Proportions and their 95% confidence intervals were computed using the Wilson score method. The analysis was performed using Python (Version 3.9).

### Experiment 2: NEJM CPC - Reasoning Quality

The models and physicians were asked to provide justifications and reasoning for their respective differential lists generated in the earlier experiment, using prompts adapted from a previous GPT-4 study (Supplement 2A). The primary outcome of this study was the quality of clinical reasoning documentation, assessed by the R-IDEA score (19). The R-IDEA is a validated 10-point scale that evaluates four key domains of clinical reasoning documentation (Supplement 2). A single specialist physician (G.L.) independently rated the reasoning documentation.

### Statistical Analysis

We compared the outputs from different models, MD-PIE, and physicians. A McNemar’s test was conducted among the models, MD-PIE, and physicians to assess the achievement of a perfect R-IDEA score. The analysis was performed in Python (version 3.9).

## Results

### Quantitative Benchmarking Results

#### Experiment 1: Zero-Shot Reasoning

In the zero-shot setting, each model attempted to provide accurate diagnoses without any specific task instructions or additional guidance. The results show that DeepSeek v3 achieved the highest accuracy of 71.6%, outperforming both OpenAI o1 (68.2%) and Gemini 2.0 Flash Thinking (66.9%) (Table 1). A possible explanation for DeepSeek’s superior performance is that its underlying architecture or training method may better generalize to unseen questions, allowing it to select the most likely diagnosis without relying on extra prompts or instructions.

**Table 1:** Zero-shot performance on the English questions of MedQA. DeepSeek v3 achieves the highest accuracy in this setting.

| Model | Accuracy (%) |
| --- | --- |
| OpenAI o1 | 68.2 |
| Gemini 2.0 Flash Thinking | 66.9 |
| DeepSeek v3 | **71.6** |

#### Experiment 2: Instruction-Guided Reasoning

Surprisingly, in this instruction-guided setting, OpenAI o1 led the performance with an accuracy of 75.7% (Table 2). When given explicit instructions on how to reason through the medical questions, OpenAI o1 led the pack with an accuracy of 75.7%. Gemini 2.0 Flash Thinking came in a close second at 74.5%, while DeepSeek v3 followed at 73.6%. The fact that OpenAI o1 improved more significantly in this instruction-guided environment suggests that it can effectively leverage structured prompts or guidelines. By contrast, Gemini 2.0 and DeepSeek, though they also improved, did not gain as much, highlighting the importance of how each model processes and integrates external instructions into its reasoning pipeline.

**Table 2:**
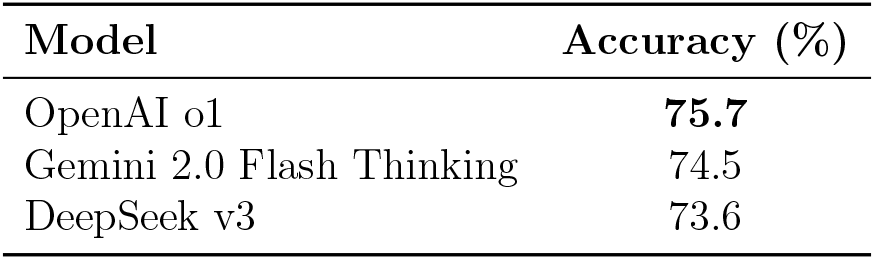
Performance on MedQA questions when a short instruction prompt is provided. OpenAI o1 performs best in this setting.

#### Experiment 3: PIE-Integrated Reasoning

In this setting, a “process of inclusion-exclusion” (PIE) method was used to iteratively narrow down a list of potential diagnoses, aiming to converge on the single most plausible answer. Here, all three models benefited from the PIE approach: DeepSeek v3 + PIE achieved the highest accuracy of 84.7%, Gemini 2.0 + PIE reached 83.2%, and OpenAI o1 + PIE attained 82.6% (Table 3). These improvements underline the value of systematically ruling in and ruling out diagnoses, suggesting that an iterative, structured reasoning framework can yield more precise outcomes regardless of the base model.

**Table 3:** Performance comparison with the PIE approach integrated. DeepSeek v3 + PIE achieves the highest accuracy at 84.7%.

| Model + PIE | Accuracy (%) |
| --- | --- |
| OpenAI o1 + PIE | 82.6 |
| Gemini 2.0 + PIE | 83.2 |
| DeepSeek v3 + PIE | **84.7** |

### Qualitative Benchmarking Results

#### Experiment 1: NEJM CPC - Differential Diagnosis Quality

We compared MD-PIE with the full version of o1, Gemini 2.0 Flash Thinking, DeepSeek V3, and specialist physicians, using clinicopathologic conferences (CPCs) from the New England Journal of Medicine (NEJM)—a recognized standard for assessing differential-diagnosis generators since the 1950s. MD-PIE, o1, and Gemini 2.0 Flash Thinking each included the correct diagnosis or a closely related helpful suggestion in their differentials 100% of the time (Bond Score: 4,5) (Figure 1). In contrast, DeepSeek V3 and specialist physicians did so in 80% of cases (Figure 1).

**Figure 1:**
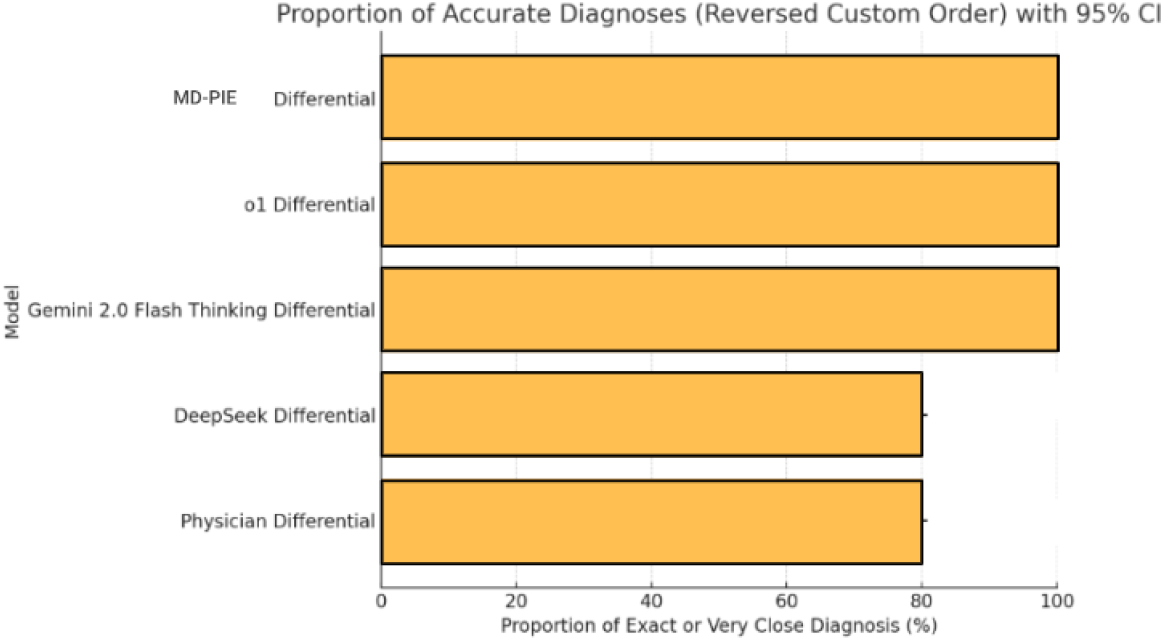
Barplot showing the accuracy of including the correct diagnosis in the differential for MD-PIE, the LLMs and physicians on the NEJM CPCs

When considering only the first suggestion in the differential list, MD-PIE’s initial choice matched the correct diagnosis in 60% of cases (o1: 40%, Physicians: 40%, Gemini 2.0 Flash Thinking: 20%, DeepSeek V3: 20%) (Figure 2). Examples of these models successfully handling complex cases can be found in Supplementary Material.

**Figure 2:**
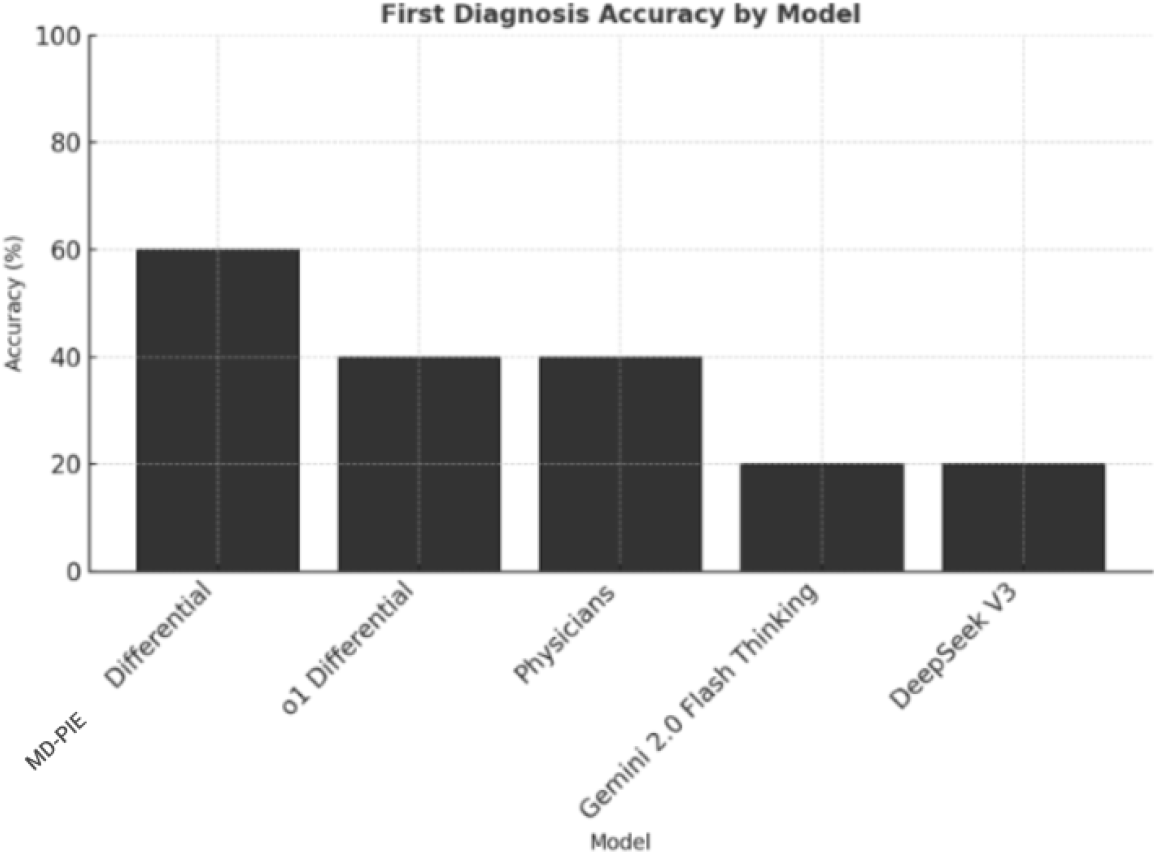
Proportion of Responses Containing the Exact or Very Close Diagnosis for MD-PIE, the LLMs and physicians on NEJM CPCs

#### Experiment 2: NEJM CPC - Reasoning Quality

We used five previously published clinical medical cases from the New England Journal of Medicine to further assess clinical reasoning, scoring each case using the Revised-IDEA (R-IDEA) system, a validated 10-point scale for evaluating four core domains of clinical reasoning. MD-PIE achieved perfect or near-perfect scores (R-IDEA 9 or 10) on all five cases, performing significantly better (P = 0.0001) than the other models and physicians (Figure 3).

**Figure 3:**
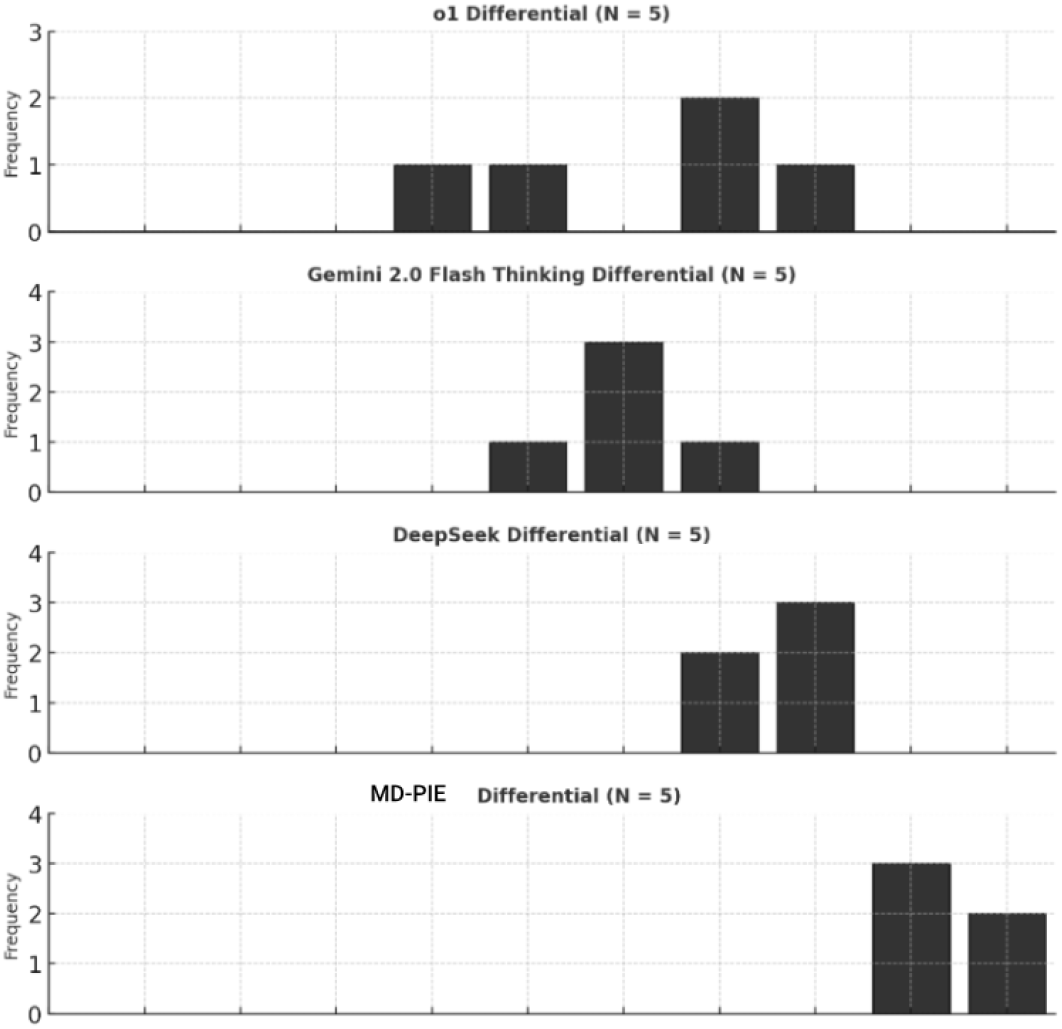
Comparison of MD-PIE, LLMs and Physicians for Clinical Diagnostic Reasoning - Distribution of R-IDEA Scores on NEJM CPCs

## Discussion

This study highlights the potential of adaptive AI systems, such as MD-PIE, in enhancing clinical diagnostic workflows by leveraging structured reasoning methodologies. By systematically narrowing the diagnostic space through iterative patient responses, MD-PIE demonstrates a capacity for advanced reasoning and transparency. The integration of a “thought” or reasoning field within its outputs ensures auditability, mitigating the risks of black-box decision-making. This transparency enables patients to share AI-driven conversational exchanges and rationales with their healthcare providers, fostering trust and facilitating the seamless incorporation of AI-generated insights into clinical decision-making (20–21).

Quantitative and qualitative benchmarking revealed significant advancements in MDPIE’s diagnostic accuracy and reasoning capabilities. MD-PIE consistently outperformed state-of-the-art models and even human benchmarks in providing high-quality diagnoses and clinical reasoning. For differential diagnosis generation, MD-PIE not only produced accurate and helpful suggestions comparable to leading models such as OpenAI o1 and Gemini

Flash Thinking but also surpassed them in identifying correct diagnoses with higher accuracy. Moreover, MD-PIE excelled in tasks requiring advanced critical thinking, such as diagnostic justification. The use of gold-standard benchmarks, such as the New England Journal of Medicine Clinicopathologic Conferences (NEJM CPCs), further underscored MD-PIE’s ability to tackle challenging diagnostic cases, highlighting its advancements in reasoning and intelligence metrics.

The results of this study also demonstrate the feasibility and impact of integrating structured reasoning approaches, such as the Problem of Inclusion-Exclusion (PIE) framework, into medical question-answering tasks. Both general-purpose and domain-specific large language models benefitted from structured query approaches, with models like DeepSeek V3 leveraging their domain-specific strengths to achieve superior performance. This indicates that combining domain-focused models with general-purpose systems could offer an optimal balance of specialization and versatility in medical reasoning. Future research may explore hybrid strategies, such as ensembling or multi-agent collaborations, to further enhance diagnostic accuracy and adaptability.

Despite these promising findings, this study has several limitations. The evaluation was limited to five cases across two experiments, restricting the generalizability of the results. Expanding the sample size to include a larger and more diverse set of cases is essential for a more comprehensive assessment of MD-PIE’s capabilities. Additionally, the study did not examine human-AI interaction, which is critical for evaluating MD-PIE’s potential to enhance clinical workflows. Future research should investigate how AI systems like MD-PIE integrate into real-world settings and interact with clinicians to optimize diagnostic processes. Furthermore, the study focused on narrow aspects of clinical reasoning, leaving areas such as surgical decision-making and interdisciplinary care unexplored. Broader studies across diverse specialties and clinical scenarios are necessary to evaluate MD-PIE’s performance comprehensively.

The adoption of AI in clinical decision-making represents an opportunity to address the significant costs—both human and financial—associated with diagnostic delays and errors (22). However, successful implementation requires the development of robust clinical frameworks to support computing infrastructure, clinician-AI interaction designs, and monitoring mechanisms (23–24). Benchmarks must be established to guide the integration of AI systems into clinical workflows, ensuring ethical and effective use. Emerging approaches, such as Medical Decision-making Agents (MDAgents), offer promising solutions by dynamically allocating tasks based on complexity, engaging individual agents for simpler cases and multidisciplinary teams for more complex scenarios. Studies of such frameworks have demonstrated improved diagnostic accuracy and reasoning, providing valuable insights into how collaborative AI systems can enhance decision-making (7–10, 22–26).

This study also underscores the need for pragmatic benchmarks that reflect real-world diagnostic challenges to foster clinician trust. Scaling experiments to include diverse cases, studying human-AI interactions in clinical workflows, and refining benchmarks will be critical for advancing the field. Ultimately, the findings presented here emphasize the transformative potential of AI systems like MD-PIE in improving diagnostic accuracy, efficiency, and overall healthcare quality. For successful adoption, it is imperative to align AI models with clinician workflows, foster trust through transparency, and ensure ethical use through robust monitoring frameworks. By advancing our understanding of how adaptive AI systems can emulate and enhance clinical reasoning, this study sets a new standard for AI-assisted medical decision-making.

## Data Availability

All data produced in the present study are available upon reasonable request to the authors

